# Debiasing and Educational Interventions in Medical Diagnosis: A Systematic Review

**DOI:** 10.1101/2022.09.12.22279750

**Authors:** Arthur Tung, Michael Melchiorre

## Abstract

**Background:** The prevalence of cognitive bias and its contribution to diagnostic errors has been documented in recent research. Debiasing interventions or educational initiatives are key in reducing the effects and prevalence of cognitive biases, contributing to the prevention of diagnostic errors. The objectives of this review were to 1) characterize common debiasing strategies implemented to reduce diagnosis-related cognitive biases, 2) report the cognitive biases targeted, and 3) determine the effectiveness of these interventions on diagnostic accuracy.

**Methods:** Searches were conducted on April 25, 2022, in MEDLINE, Embase, Healthstar, and PsycInfo. Studies were included if they presented a debiasing intervention which aimed to improve diagnostic accuracy. The Rayyan review software was used for screening. Quality assessments were conducted using the JBI Critical Appraisal Tools. Extraction, quality assessment, and analysis were recorded in Excel.

**Results:** Searches resulted in 2232 studies. 17 studies were included in the final analysis. Three major debiasing interventions were identified: tool use, education of biases, and education of debiasing strategies. All intervention types reported mixed results. Common biases targeted include confirmation, availability, and search satisfying bias.

**Conclusion:** While all three major debiasing interventions identified demonstrate some effectiveness in improving diagnostic accuracy, included studies reported mixed results when implemented. Furthermore, no studies examined decision-making in a clinical setting, and no studies reported long-term follow-up. Future research should look to identify why some interventions demonstrate low effectiveness, the conditions which enable high effectiveness, and effectiveness in environments beyond vignettes and among attending physicians.

**PROSPERO registration number:** CRD42022331128

## Introduction

Diagnostic error has long been recognized as a significant contributor to patient harm. Missed diagnoses or incorrect diagnoses are surprisingly common, with average diagnostic error rates hovering around 13–15% under both usual practice and research-directed conditions (Newman-Toker et al., 2021; Singh, 2013). These rates generally agree with autopsy studies which report that 10–20% of cases show major diagnostic discrepancies (Graber, 2013). Diagnostic errors are not only not uncommon but also dangerous; it has been demonstrated that misdiagnosis accounted for the largest fraction of severe patient injury in the United States and was twice as likely to result in patient death or disability than other error categories (Saber Tehrani et al., 2013). Moreover, this does not account for less severe, but still consequential, effects of misdiagnosis for patients, including decreased quality of life, further disease progression, and unnecessary costs of ineffective treatments.

Previous literature focusing on cognitive psychology as it relates to medicine has found that cognitive biases and errors are major factors in generating diagnostic errors (Saposnik et al., 2016). Qualitative studies have found that cognitive errors accounted for 32% of all misdiagnoses in internal medicine settings (Schiff et al., 2009), and contributed to up to 92% of all diagnostic errors in emergency medicine settings (Okafor et al., 2016). Based on a retrospective analysis of identified and reported diagnostic discrepancies, Graber and colleagues estimated that cognitive factors contributed to 74% of misdiagnoses, averaging 4.32 cognitive contributions to error per case reviewed (320 cognitive errors in 74 cases). Due to the contribution of cognitive factors to diagnostic error and the high prevalence of diagnostic error, research, identification, and implementation of effective debiasing interventions and educational initiatives which aim to decrease common cognitive pitfalls in clinical reasoning are vital to improving diagnostic accuracy and patient outcomes.

Dual-process theory, as initially proposed by William James and further developed by Daniel Kahneman and Amos Tversky, has been adapted from the social psychology field to explain how clinician cognitive errors could arise from the use of heuristics (Klein, 2008). System 1 or type 1 processing is the mode of reasoning best described as fast, intuitive, automatic, and heuristics based. Type 1 is also the “default” mode of thinking. Cognitive psychologists estimate most spend around 95% of their time in type 1 (Lakoff & Johnson, 1999). In contrast, type 2 processing is slow, conscious, analytical, and controlled (Kahneman, 2003).

Type 1 processing has been associated with the generation of cognitive biases when the automatic and unconscious use of heuristics is inappropriate for the situation (Norman et al., 2017). However, it must be noted that the use of type 1 processing and heuristics is usually appropriate and leads to the correct responses. Furthermore, type 2 thinking may still lead to incorrect responses if applied inappropriately (Evans & Stanovich, 2013). The cause of cognitive biases is the result of failures of both type 1 and 2 processes, type 1 for generating the error and type 2 for failing to detect and correct the error (Norman et al., 2017). Therefore, interventions which seek to minimize the effects and prevalence of cognitive biases should look to further develop the clinician’s ability to recognize unchecked use of heuristics and rely more on type 2 thinking when appropriate or teach debiasing strategies to clinicians. Type 2 thinking could also be encouraged or forced with the use of planned diagnostic slowdowns or timeouts (Yale et al., 2022). Alternatively, interventions could look to implement standardized approaches to diagnosis through the assistance of structured checklists or protocols.

In recent decades, medical educators have developed and implemented various debiasing interventions and educational initiatives within the medical curriculum and graduate medical education programs. In 2013, Croskerry and colleagues categorized various published and theorized strategies for cognitive debiasing (Croskerry et al., 2013), including the education of cognitive biases, implementation of decision-making support tools and protocols (i.e. checklists, diagnostic timeouts, and reminder systems), education of debiasing strategies (i.e. metacognition and consider-the-opposite strategy), and workplace improvements (i.e. avoidance of fatigue and sleep deprivation, and group decision strategy). Since 2013, educators have attempted to implement some of the strategies presented above in usual practice and in undergraduate or graduate medical education curriculums. However, no systematic review to our knowledge has yet to examine the effectiveness of these recent strategies in improving diagnostic accuracy or factors supporting sound clinical decision-making.

A review of the literature which reports on the effectiveness of known interventions in reducing specific cognitive biases would help guide the development and implementation of future strategies. Given the status of the research landscape outlined above, the purpose of this systematic review was to 1) characterize and categorize the most common debiasing strategies implemented to reduce diagnosis-related cognitive biases in the literature, 2) report the types of cognitive biases targeted by each intervention, and 3) determine the effectiveness these interventions have on diagnostic accuracy.

## Methods

This systematic review was conducted and reported according to the updated Preferred Reporting Items for Systematic Reviews and Meta-Analyses (PRISMA) 2020 statement (Page et al., 2021). The review’s protocol was registered with PROSPERO under the registration number CRD42022331128.

### Literature Search

Searches were conducted on April 15, 2022, through four electronic databases (MEDLINE, Embase, PsycINFO, and Healthstar) using the Firefox browser. Search results were exported in Research Information Systems (RIS) format and imported to Rayyan for Systematic Reviews (Ouzzani et al., 2016). See supplementary materials for the full search strategy.

### Inclusion and Exclusion Criteria

Studies were included if they 1) reported the implementation of at least one intervention or educational initiative related to diagnosis, 2) focused on reducing the prevalence or effect of cognitive biases in medical diagnosis, 3) examined diagnostic decision-making in a clinical setting or hypothetical clinical setting (i.e., patient vignettes), 4) participants in the intervention included medical students, resident physicians, attending physicians, physician assistants (PA), nurse practitioners, (NP), PA students, or NP students, and 5) were classified as an original study or conference abstract. Studies were excluded if they 1) did not explicitly ascribe at least one cognitive bias to diagnostic error, 2) were classified as a letter, review, commentary, editorial, book, or protocol, or 3) only suggested potential debiasing strategies without implementing at least one debiasing intervention.

Two screeners independently reviewed all studies using the Rayyan software program through one round of title and abstract screening and one round of full-text review. Screeners were blinded to each other’s decisions until the end of each screening round, after which a discussion was held to resolve any conflicts.

### Data Extraction

Two reviewers independently extracted the included studies and imported them into a preformatted Excel worksheet. Information extracted included: author names and publication year, medical specialty, number of participants and their medical experience, intervention type and nature, cognitive bias targeted, and debiasing intervention efficacy.

### Quality Assessment

Internal validity and quality of included studies were assessed with JBI’s “Checklist for Randomized Control Trials” or “Checklist for Quasi-Experimental Studies (Non-Randomized Experimental Studies)” tool. Additionally, each study was given a subjective rating of poor, fair, or good to describe the raters’ overall appraisal.

## Results

### Search Results

Database searches yielded 2232 articles, 1166 of which were identified as unique by the Rayyan software program. 1142 articles were excluded during the title and abstract screening. Inter-rater reliability (IRR, average agreement) during the abstract screening was 98.88%, with a Cohen’s kappa coefficient (κ) of 0.73, indicating substantial agreement. The remaining 24 articles were retrieved and underwent full-text screening, during which 7 additional articles were excluded. IRR in this round was 86.96%, with a κ of 0.59, indicating moderate agreement. 17 articles were included in the final analysis.

See figure 1 for PRISMA article selection information.

**Figure 1.**
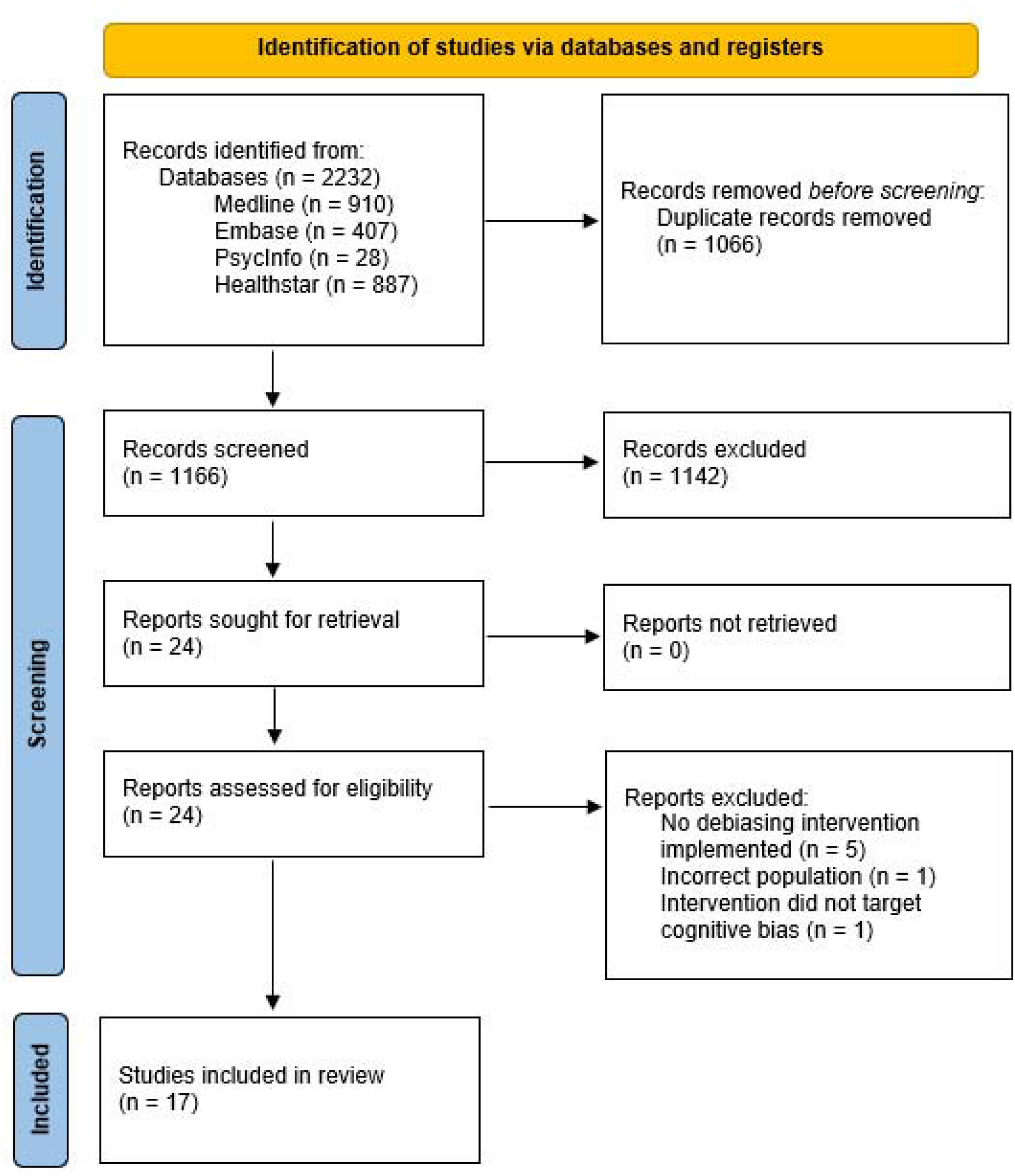
PRISMA flowchart of selected studies

### Study Characteristics

The final analysis included a total of 1530 participants across 17 studies: 849 medical students (849/1530, 55.49%), 485 residents (465/1530, 30.39%), 30 attending physicians (30/1530, 1.96%), and 186 other healthcare professionals (186/1530, 12.16%). 11 of the studies were randomized controlled trials (RCT) (11/17, 64.71%), and 6 were quasi-experimental studies (QES) (6/17, 35.29%). Among the 17 studies, 13 examined diagnostic accuracy in a hypothetical clinical setting via case vignettes (13/17, 76.47%), 2 examined knowledge of reasoning/diagnostic skills via assessments involving patient vignettes (2/17, 11.76%), 1 examined bias recognition ability in pre-diagnosed patient vignettes (1/17, 5.88%), and 1 examined attitudes towards a conference (1/17, 5.88%). Only one of the included studies involved an actual clinical aspect (1/17, 5.88%), but did not evaluate diagnostic decision-making in an actual clinical environment.

### Data Quality

IRR in quality assessment was 76.5% (13/17). 4 studies were rated as poor (4/17, 23.53%), 6 studies were rated as fair (6/17, 35.29%), and 7 studies were rated as good (7/17, 41.18%). The majority of studies rated good were successful interventions; out of 7, 5 were successful (5/7, 71.43%). The inverse was observed in studies rated poor, in which the majority were unsuccessful; out of 4, 1 was successful. (1/4, 25%).

See supplementary materials for quality assessment details (McGuinness & Higgins, 2021).

### Objective 1: Debiasing Strategies Implemented

Three broad categories of debiasing intervention types were identified during the analysis of the included studies. The distribution of them across the included studies was as follows: 8 of the studies aimed to increase awareness/education of a bias (8/17, 47.06%), 7 aimed to provide education of debiasing strategies (7/17, 41.18%), and 6 used a tool such as a debiasing checklist (6/17, 35.29%). 4 of the studies’ debiasing intervention utilized more than one of the three intervention types (4/17, 23.53%), but none of the studies used all three (0/17, 0%). Among the three intervention types, several subcategories were found, including classes, workshops, checklists, metacognitive strategies, and long-term educational curricula.

Refer to Table 1 for full details of the nature of implemented interventions and their effectiveness.

**Table 1.**
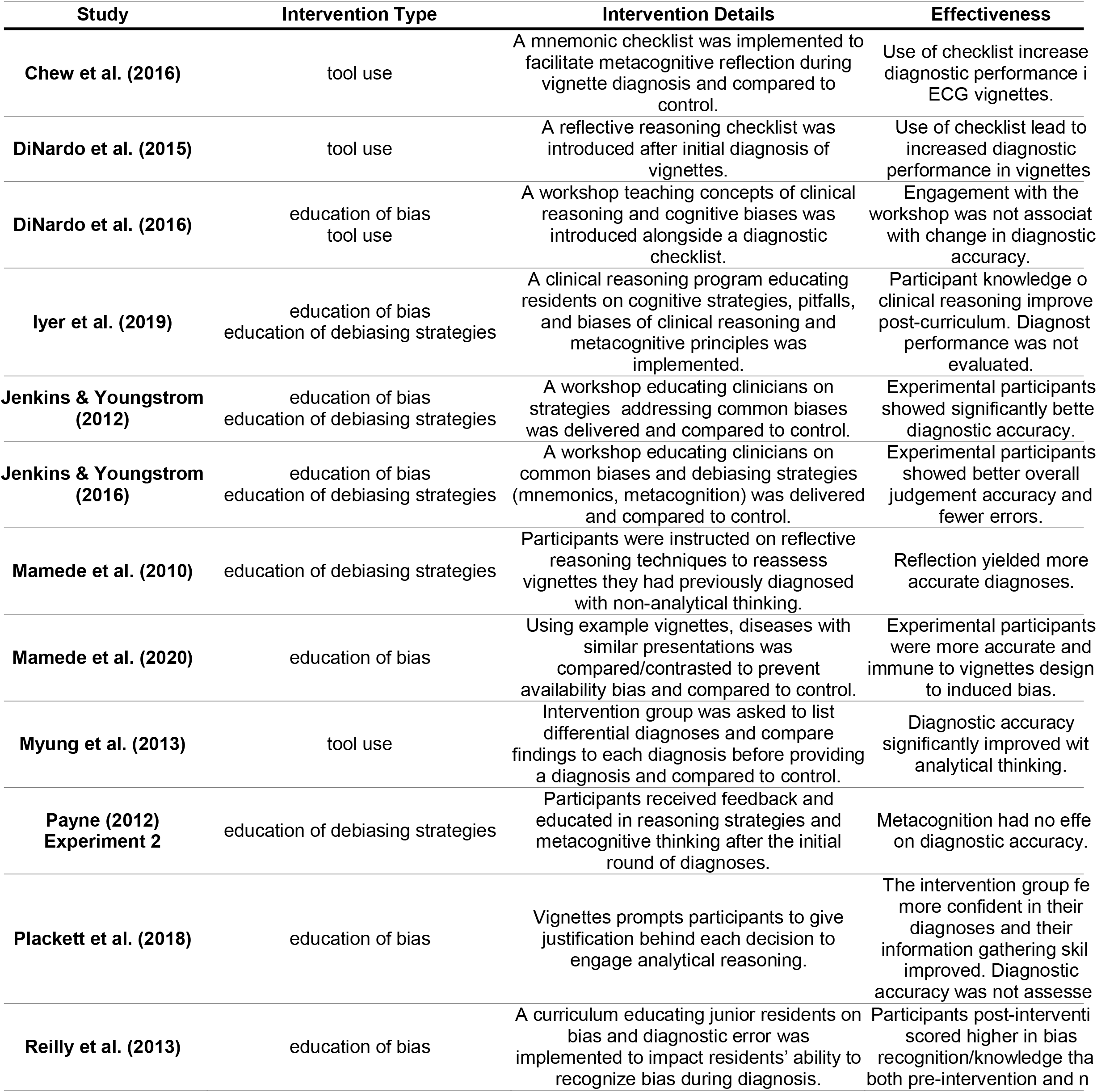
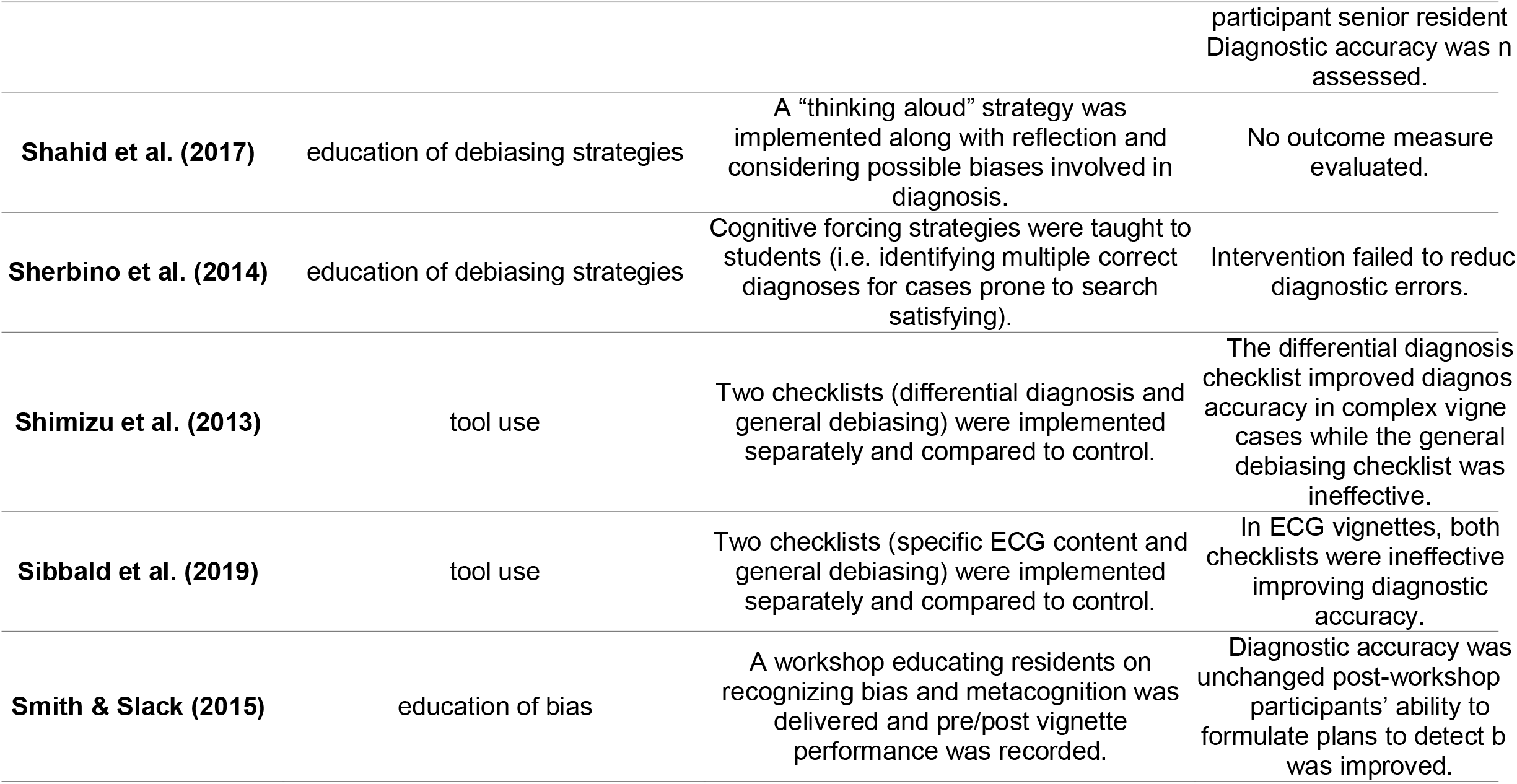
Details and effectiveness of debiasing interventions

### Objective 2: Cognitive Biases Targeted

8 different biases were targeted across the included studies. The three most reported biases, excluding general/non-specific cognitive bias, were as follows: availability bias, targeted in 5 studies (5/17, 29.41%); search satisfying, targeted in 4 studies (4/17, 23.53%); and confirmation bias, targeted in 3 studies (3/17, 17.65%). 9 studies (9/17, 52.94%) did not target specific biases but biases in general. Metacognition, whether by education of debiasing strategy or through the implementation of checklists designed to induce reflective thinking, was found to more commonly counteract availability bias, confirmation bias, and search satisfying. No trends were found regarding any other specific bias being commonly treated with any specific debiasing intervention.

See Table 2 for identified cognitive biases and other cognitive biases mentioned in this review (Croskerry, 2003).

**Table 2.**
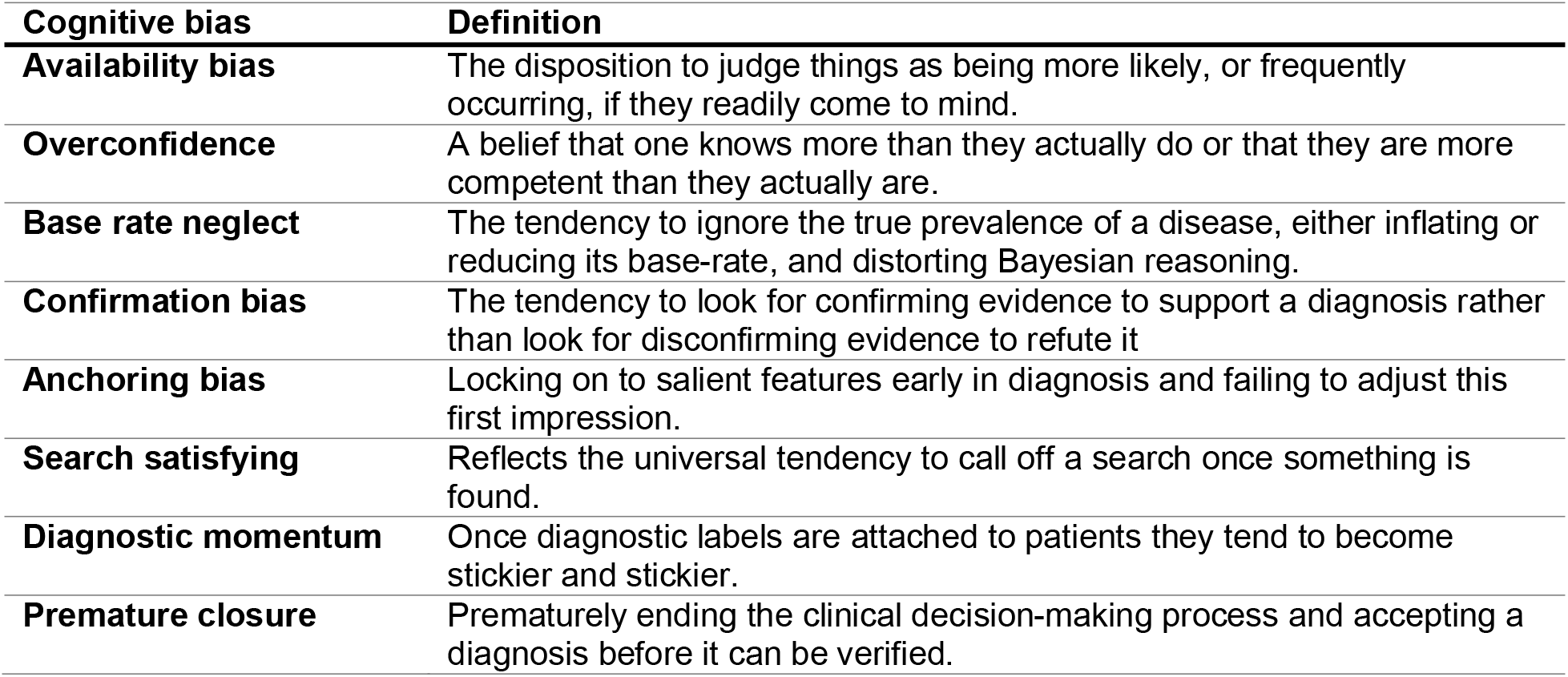
Definitions of cognitive biases mentioned

| **Cognitive bias** | **Definition** |
| --- | --- |
| **Availability bias** | The disposition to judge things as being more likely, or frequently occurring, if they readily come to mind. |
| **Overconfidence** | A belief that one knows more than they actually do or that they are more competent than they actually are. |
| **Base rate neglect** | The tendency to ignore the true prevalence of a disease, either inflating or reducing its base-rate, and distorting Bayesian reasoning. |
| **Confirmation bias** | The tendency to look for confirming evidence to support a diagnosis rather than look for disconfirming evidence to refute it |
| **Anchoring bias** | Locking on to salient features early in diagnosis and failing to adjust this first impression. |
| **Search satisfying** | Reflects the universal tendency to call off a search once something is found. |
| **Diagnostic momentum** | Once diagnostic labels are attached to patients they tend to become stickier and stickier. |
| **Premature closure** | Prematurely ending the clinical decision-making process and accepting a diagnosis before it can be verified. |

### Objective 3: Effectiveness of Interventions

Of the 17 studies, 8 reported an increase in diagnostic accuracy attributed to at least one debiasing intervention (8/17, 47.06%), and 5 of the studies reported no improvement in diagnostic accuracy post-intervention (5/17, 29.41%). 4 studies reported improvements in areas other than diagnostic accuracy, such as knowledge of cognitive biases and potential debiasing strategies (4/17, 23.53%). Note that studies could implement more than one type of intervention. The success rates of each of the three intervention types were as follows: among interventions aiming to increase awareness/education of bias, 6 of the 8 studies reported successful improvement of diagnostic accuracy (6/8, 75%) and 2 reported ineffectiveness (2/8, 25%); among interventions aimed at providing education of debiasing strategies, 4 of the 7 reported successful improvement of diagnostic accuracy (4/7, 57.14%), 2 reported ineffectiveness (2/7, 28.57%), and 1 study did not report effectiveness (1/7, 14.29%); and among interventions involving tool use, 4 of the 6 studies reported successful improvement of diagnostic accuracy (4/6, 66.67%) including 1 study which reported one effective checklist and one ineffective checklist, and 2 reported ineffectiveness (2/6, 33.33%). Of the 4 studies that did not directly aim to improve diagnostic accuracy (4/17, 23.53%), 2 of the studies focused on measuring participants’ knowledge of cognitive/diagnostic error (2/4, 50%), 1 study examined the information-gathering ability of participants (1/4, 25%), and one study did not have an outcome measure (1/4, 25%).

## Discussion

Across 17 studies included in the final analysis, three broad categories of intervention types were identified. These categories are: 1) education or increase awareness of cognitive biases, 2) education of debiasing strategies, and 3) implementation of decision-making support tools.

### Characterizing identified debiasing interventions

#### Education or increase awareness of cognitive biases

In 2013, Croskerry and colleagues characterized the average clinician’s relationship with cognitive bias in medical decision-making at the “precontemplative level” (Croskerry et al., 2013). The precontemplative stage is denoted by general unawareness of the powerful influences of cognitive biases on decision-making despite understanding how to make correct decisions (Prochaska et al., 1992). The education of biases, therefore, provides the prerequisite knowledge needed to identify instances of cognitive biases and initiate debiasing. This type of intervention assumes that knowledge of biases will translate directly into increased proficiency in identifying those biases outside of education-driven environments. However, previous interventional studies have either failed to report the effects of intervention on diagnostic accuracy (Bond et al., 2004; Ogdie et al., 2012) or reported limited intervention effectiveness in non-clinical environments (Sherbino et al., 2011).

In the present review, 8 studies utilized education of biases as one of their implemented interventions. Most studies used conventional forms of education, namely lectures (Iyer et al., 2019; Jenkins & Youngstrom, 2016; Reilly et al., 2013; Smith & Slack, 2015), group discussion (Reilly et al., 2013), workshops (DiNardo et al., 2016; Jenkins & Youngstrom, 2012), and reading exercises on the role of bias in diagnosis (Smith & Slack, 2015). Education of biases was also often combined with other types of interventions (i.e., debiasing checklists) during workshops and educational initiatives (Smith & Slack, 2015). Other studies provided firsthand experience of cognitive biases via vignettes designed to induce bias (Mamede et al., 2020; Plackett et al., 2020; Reilly et al., 2013). One such intervention reported by Mamede and colleagues indirectly increased physician awareness of availability bias by focusing on discriminatory elements between diseases with similar presentations (Mamede et al., 2020). Therefore, this method may demonstrate that education in the process of reasoning, like other studies of this type of intervention, still plays a secondary role to education of content knowledge in preventing bias-induced diagnostic error. Though studies which do focus on educating clinicians in the process of reasoning have similarly produced positive results (Iyer et al., 2019; Jenkins & Youngstrom, 2016; Jenkins & Youngstrom, 2012). Furthermore, the study population were residents who may have benefited most from knowledge of diseases rather than knowledge of reasoning processes.

#### Education of debiasing strategies

Metacognition has long been suggested as the principle debiasing strategy for tackling cognitive errors in diagnosis (Croskerry, 2003). Metacognition is a reflective approach to decision-making which examines the reasoning process used to arrive at an initial judgement (thinking about thinking) (Graber, 2003). The act of reflection allows for decoupling from the intuitive type 1 process and engaging type 2 thinking to verify initial judgements by checking for conflicting evidence and alternatives (Croskerry et al., 2013). Education of other strategies, namely group decision strategy and exposure control (i.e., limiting exposure to knowledge of another clinician’s working diagnosis before forming an initial impression), may also encourage reflective reasoning, though articles investigating these interventions are limited. In addition, group decision strategy alone may succumb to diagnostic momentum if the initial diagnosis is not critically examined (O’Sullivan & Schofield, 2018). 7 studies were identified as using education of debiasing strategies in their interventions. Unsurprisingly, most studies (6/7, 85.7%) within this type of intervention utilized metacognitive principles in some capacity (Iyer et al., 2019; Jenkins & Youngstrom, 2016; Mamede et al., 2010; Payne, 2012; Shahid et al., 2017; Sherbino et al., 2014). One study did not specify the exact debiasing strategy implemented (Jenkins & Youngstrom, 2012). One approach to implementing metacognition was “think aloud” exercises where participants explain the rationale of tests and diagnoses, allowing for reflective reasoning and engagement with type 2 processing (Shahid et al., 2017). Another popular approach was consider-the-alternative strategies, which encouraged critical examination of the differential diagnosis and seeking evidence which opposed the leading diagnosis (Jenkins & Youngstrom, 2016; Mamede et al., 2010). While these approaches should theoretically engage analytical thinking and encourage the detection of cognitive biases, it is unclear if participants would call upon these methods in actual clinical situations.

#### Decision-making support tools

Checklists are commonly used in other industries which are susceptible to human error and cognitive biases, namely in aviation (Gawande, 2009). Though many components of checklists in high-risk industries are obvious and familiar, the routine use of them ensures a reproducible approach across all users and could prevent easily avoided disasters (Ely et al., 2011). Surgical safety checklists are one example of how the healthcare sector has successfully adopted widespread use of checklist-based interventions to reduce human errors (Armstrong et al., 2022). In medical diagnosis, different checklists can be used in different steps of diagnosis. For example, knowledge retrieval checklists can be used to ensure information gathered on patients are unbiased, differential diagnosis checklists enforce consider-the-alternative strategies when considering the leading diagnosis, and general debiasing checklists allow for reflective thinking of the reasoning process (Ely et al., 2011). Other decision-making support tools have been developed but were not discussed in the review because they do not explicitly debias. For example, computer-based support tools like DXplain suggest possible diagnoses which fit with patient symptoms and may help complete the differential diagnosis (Barnett et al., 1987). 6 studies were found to have used checklists as one of their implemented interventions, with multiple studies implementing multiple checklists. Most studies (5/6, 83.3%) reported using a general debiasing checklist (Chew et al., 2016; DiNardo et al., 2015, 2016; Shimizu et al., 2013; Sibbald et al., 2019), which encourages metacognition and is sometimes used in conjunction with the education of metacognitive principles (Chew et al., 2016). One study only used a differential diagnosis checklist (Myung et al., 2013). In one approach, Chew and colleagues proposed a general debiasing checklist mnemonic in an attempt to enable clinicians to conveniently apply the checklists in clinical situations, arguing that the use of a more robust checklist would be cumbersome to use outside of educational environments (Chew et al., 2016).

Though the practicality of these checklists was questioned by Chew and other authors, especially in simpler clinical cases (Shimizu et al., 2013), only one study attempted to assess the usability of any checklists in a clinical setting and found that the checklist was seldomly used despite favourable feedback from clinician participants (DiNardo et al., 2016).

### Cognitive biases targeted by interventions

The most targeted cognitive biases were availability bias, search satisfying, and confirmation bias. All three biases are known to cause fixation on one leading diagnosis, which may lead to premature closure (Croskerry, 2003). Among the included articles, metacognitive principles delivered in the form of an educational intervention and/or checklist were shown to have been used to target all three biases in almost all instances. This is unsurprising, as metacognition encourages clinicians to reconsider the working hypothesis by analyzing the reasoning steps taken to arrive at that conclusion, which would help alleviate a fixation on one diagnosis. This finding, however, could also be the result of reporting bias.

Most interventions analyzed did not target specific cognitive biases or targeted a wide variety of biases. This might be a result of researchers attempting to develop solutions which can be applied to a wide array of situations, thus encouraging non-specific targeting of biases.

### Ineffectiveness of some described interventions

5 studies reported at least one ineffective intervention, which collectively includes interventions from all three types of interventions discussed thus far. The authors’ suggested reasons for the negative results vary greatly between studies and are largely speculative, but nevertheless may provide valuable insight into what future studies must avoid in designing their interventions.

Shimizu and colleagues reported one ineffective checklist intervention where their medical student participants unsuccessfully utilized a general debiasing checklist developed by Ely and colleagues (Ely et al., 2011; Shimizu et al., 2013). The authors suggested that due to the limited clinical experience that the participants had, heuristic thinking among the students may have been sparse. Therefore, a debiasing checklist would not have affected performance as cognitive bias occurs due to a fault in heuristics usage (Committee on Diagnostic Error in Health Care, 2015). Thus, if a debiasing intervention were to be tested on participants with relatively limited experience, such as medical students or junior residents, researchers should first ensure that participants demonstrate the use of heuristics or evidence of cognitive bias in the clinical situations they intend on examining.

The same debiasing checklist was also reported to be ineffective by Sibbald and colleagues (Sibbald et al., 2019). However, due to the use of simpler case vignettes, the baseline performance of both novices and experts was higher than in previous studies, which may explain the small effect size (Sibbald et al., 2013). Furthermore, a previously validated content-specific checklist also failed to demonstrate significant results in the same study (Sibbald et al., 2013, 2014), which could suggest a flaw in the study methodology which caused both checklists to be ineffective. Alternatively, since the studies examined diagnosis specifically in ECG, these results could show that debiasing checklists designed for general use are ineffective in specialized environments. High pre-intervention performance was also a suggested reason for the low effectiveness of another intervention implemented by Smith and Slack. The educational workshop demonstrated a low effect size, which was speculated to be due to the inconsistent assessment pre-workshop by attending physicians who were unfamiliar with the study evaluations.

Other instances of low intervention effectiveness include a debiasing checklist intervention reported by DiNardo and colleagues and a metacognitive education intervention by Payne. DiNardo noted highly variable use of the implemented tool and that it was used mostly by non-participant clinicians (i.e., attending physicians rather than resident participants). These factors may have contributed to the not significant findings (DiNardo et al., 2016). Payne reported that participants were distracted by graphical representations of mental models included in the educational intervention and did not spend a great deal of time on the metacognitive feedback (Payne, 2012).

### Concerns with current interventions and future research

While all three broad debiasing intervention types discussed demonstrated some promise in improving diagnostic accuracy, there are several problems which paint an unclear picture of their effectiveness in practice.

Most significantly, no intervention was evaluated in real clinical contexts. Thus, the useability and actual effectiveness of each intervention is unclear. Furthermore, all three debiasing interventions reported some instances of ineffectiveness; the exact reasonings for ineffectiveness are heterogeneous. Studies also examined the effectiveness of interventions immediately or soon after delivery of interventions. This experimental choice is logical for the purposes of research but puts the long-term effectiveness of interventions into question. Lastly, while it is immediately obvious why nearly all participants in debiasing interventions and initiatives are medical students and resident physicians, it is known that attending physicians are also vulnerable to cognitive bias (Croskerry, 2003). Therefore, the reported effectiveness of each intervention may not be accurate in attending physician populations. Moreover, a lack of attending physician participants in the included studies suggests a troubling lack of focus on the effects of cognitive biases in continued medical education.

Due to these flaws in the interventions examined in this review, we recommend that future studies should further examine the effectiveness of each intervention in 1) real clinical practice, 2) with participant populations which include attending physicians and other practicing clinicians responsible for diagnosis (i.e., NP and PA), 3) report the long term retention of educational materials or intervention usage, and 4) investigate reasoning behind ineffectiveness and low intervention usage. Future studies should also examine whether effectiveness and intervention usage in case vignettes translate directly to clinical practice.

### Limitations

There are limitations to this review which should be acknowledged. First, articles were only included if the intervention specifically mentions that it is a debiasing intervention or reduces the negative impacts of cognitive biases in diagnostic decision-making. This criterion meant that some articles that may also reduce the negative impacts of cognitive biases were not included as they did not explicitly identify themselves as such. Second, due to the heterogeneity of the extracted data, we were unable to provide a formal meta-analysis. While we were able to categorize interventions into three different types, the details of each intervention and outcome measures between studies varied greatly. Third, while the inclusion criteria specified the inclusion of NP, PA, and attending physician participants, no NP and PA participants were included in the final analysis and a limited number of attending physician participants were included. Fourth, the scope of this review was limited by the number of databases chosen. A more comprehensive search could have led to more reliable and robust findings. Lastly, all studies analyzed decision-making in a vignette context which may not accurately reflect real-world practice.

## Conclusion

In this review, we characterized and evaluated implemented debiasing interventions and educational initiatives on cognitive biases in diagnosis. Though the strategies discussed in this review reveal great potential in their effects on improving diagnostic accuracy, further research is required to develop more effective and practical solutions in clinical environments. We advocate for future research on the viability and useability of debiasing strategies in real clinical practice among practising physicians as well as students and resident physicians. In addition, researchers and medical educators should account for the time and cognitive restraints of real clinical practice and implement feedback from clinician participants to increase the useability of implemented debiasing strategies. Due to the prevalence and consequences of cognitive biases on diagnostic error, successful debiasing interventions have the potential to significantly reduce medical errors and improve patient outcomes (Graber et al., 2005; Okafor et al., 2016; Saber Tehrani et al., 2013).

## Data Availability

All data produced in the present study are available upon reasonable request to the authors

## Author Contributions

Both authors conceptualized the review, drafted the study protocol on PROSPERO, screened collected articles, extracted data, conducted quality assessment, and contributed equally to the creation of all figures and tables. AT formulated and executed the search strategy, conducted the formal analysis, drafted the abstract, introduction, discussion, and cover letter. MM drafted the methods and results, managed all citations, and revised all written materials. All data used and synthesized in this review are available from the corresponding author (AT) upon request.

## Acknowledgements

The authors would like to thank Samantha Villamor for her assistance in the manuscript editing process.

