## Supplementary Materials for "Debiasing and Educational Interventions in Medical Diagnosis: A Systematic Review"

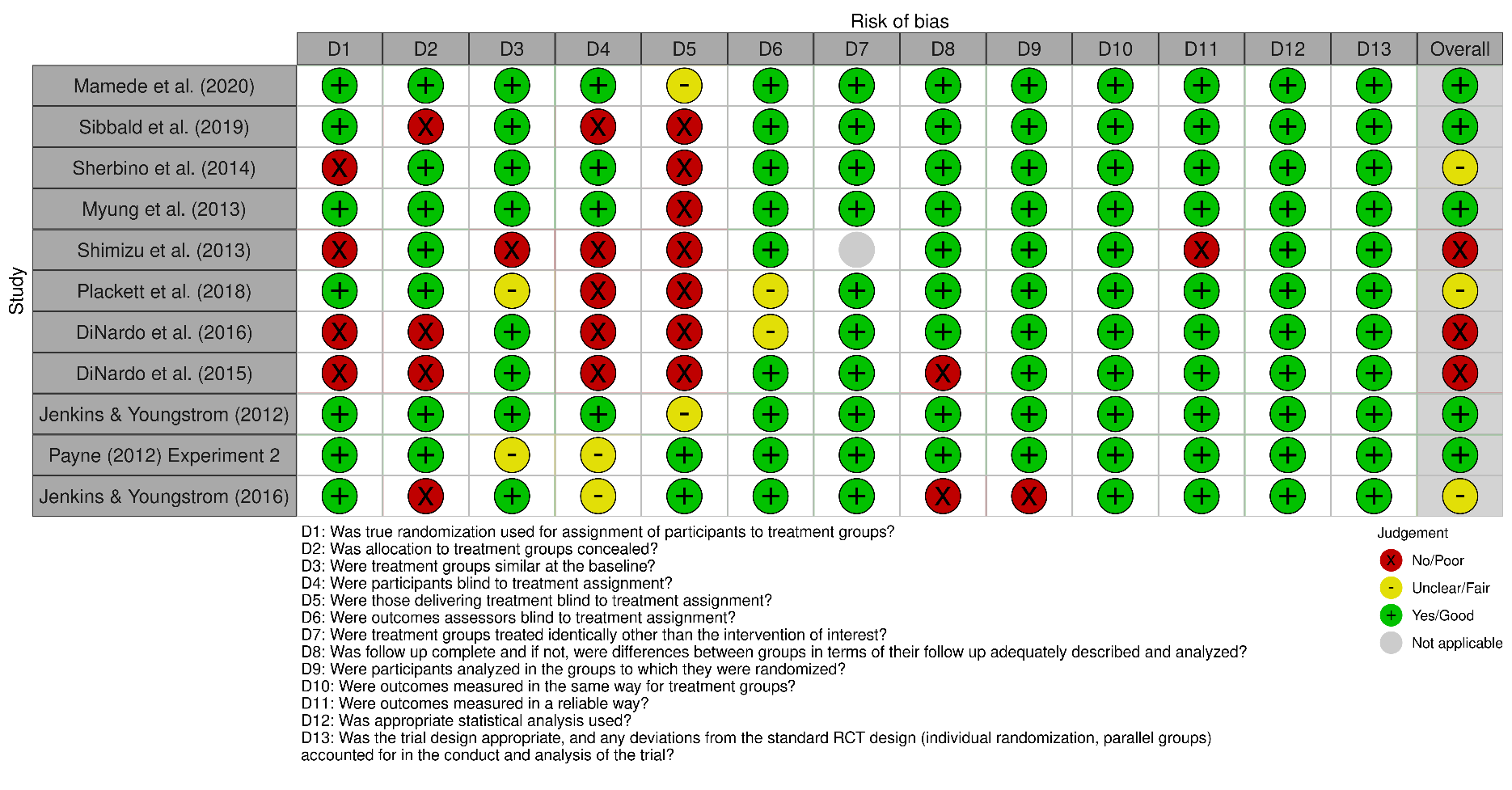
Figure S1. Randomized control trials risk of bias assessments

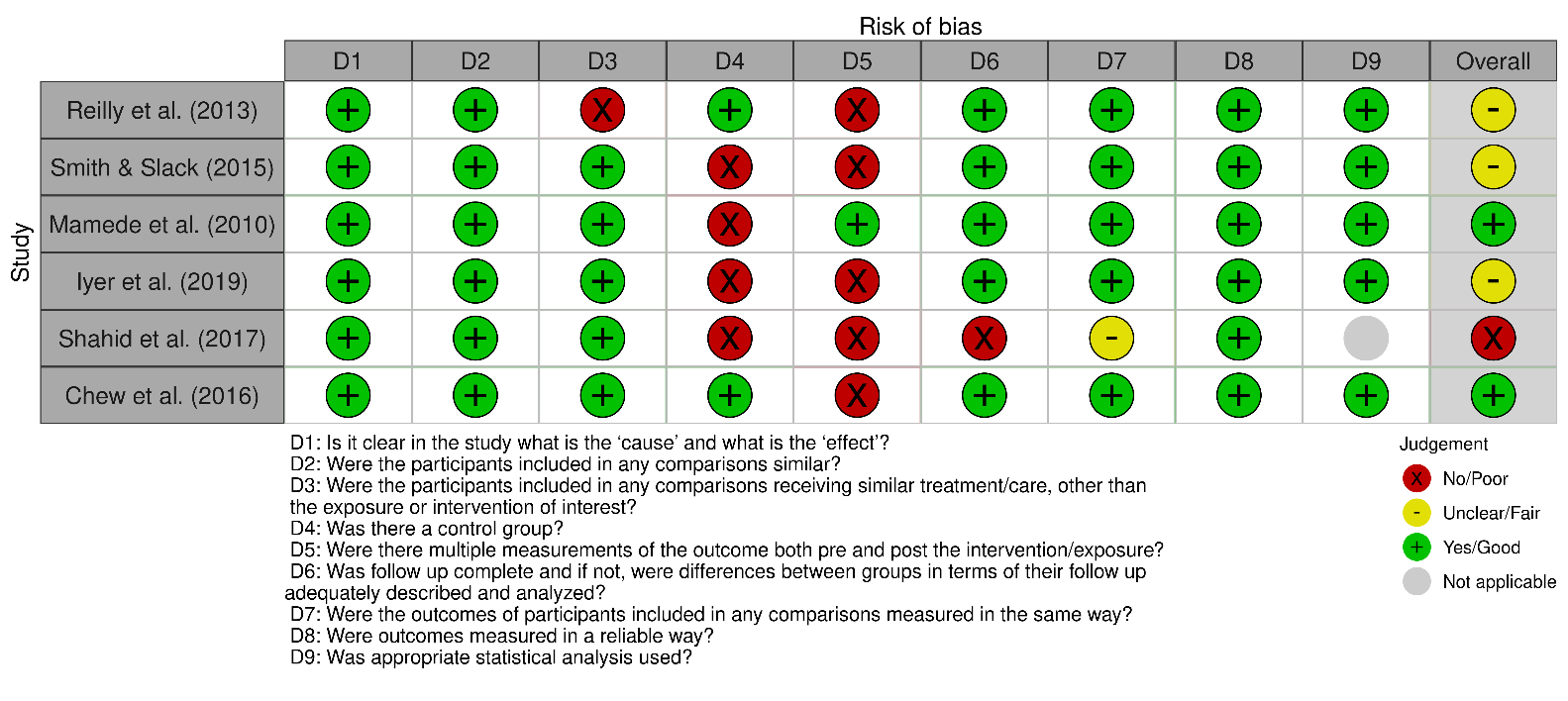
Figure S2. Quasi-experimental studies risk of bias assessments

Search Strategy

Ovid MEDLINE: Epub Ahead of Print, In-Process & Other Non-Indexed Citations, Ovid MEDLINE® Daily and Ovid MEDLINE® <1946-Present>

1	Cognitive bias.mp.	1744

2	premature closure.mp.	814

3	exp Heuristics/ or heuristics.mp.	4117

4	exp Bias/ or anchoring bias.mp.	73700

5	overconfidence.mp.	608

6	fallacy.mp.	1797

7	exp Prejudice/ or prejudice.mp.	37695

8	1 or 2 or 3 or 4 or 5 or 6 or 7	119367

9	diagnostic error.mp. or exp Diagnostic Errors/	122423

10	misdiagnosis.mp.	17068

11	exp Delayed Diagnosis/ or exp Diagnosis/ or diagnosis.mp. or exp Missed Diagnosis/	10681337

12	9 or 10 or 11	10683801

13	8 and 12	61328

14	debias.mp.	30

15	debiasing strategy.mp.	10

16	medical education.mp. or exp Education, Medical/	200243

17	educational initiative.mp.	385

18	bias mitigation.mp.	30

19	debiasing.mp.	193

20	14 or 15 or 16 or 17 or 18 or 19	200742

21	13 and 20	959

22	limit 21 to ("review" or "scientific integrity review" or "systematic review")	49

23	21 not 22	910

Embase Classic+Embase <1947 to 2022 April 22>

1	Cognitive bias.mp.	4002

2	premature closure.mp.	1335

3	exp Heuristics/ or heuristics.mp.	5142

4	exp Bias/ or anchoring bias.mp.	37939

5	overconfidence.mp.	716

6	fallacy.mp.	2256

7	exp Prejudice/ or prejudice.mp.	8895

8	1 or 2 or 3 or 4 or 5 or 6 or 7	59454

9	diagnostic error.mp. or exp Diagnostic Errors/	117433

10	misdiagnosis.mp.	25135

11	exp Delayed Diagnosis/ or exp Diagnosis/ or diagnosis.mp. or exp Missed Diagnosis/	9923600

12	9 or 10 or 11	9925150

13	8 and 12	22435

14	debias.mp.	25

15	debiasing strategy.mp.	11

16	medical education.mp. or exp Education, Medical/	381660

17	educational initiative.mp.	692

18	bias mitigation.mp.	27

19	debiasing.mp.	212

20	14 or 15 or 16 or 17 or 18 or 19	382401

21	13 and 20	453

22	limit 21 to ("review" or "scientific integrity review" or "systematic review")	46

23	21 not 22	407

APA PsycInfo <1806 to April Week 3 2022>

1	Cognitive bias.mp.	5317

2	premature closure.mp.	133

3	exp Heuristics/ or heuristics.mp.	7026

4	exp Bias/ or anchoring bias.mp.	66

5	overconfidence.mp.	1316

6	fallacy.mp.	2277

7	exp Prejudice/ or prejudice.mp.	22661

8	1 or 2 or 3 or 4 or 5 or 6 or 7	38054

9	diagnostic error.mp. or exp Diagnostic Errors/	182

10	misdiagnosis.mp.	2304

11	exp Delayed Diagnosis/ or exp Diagnosis/ or diagnosis.mp. or exp Missed Diagnosis/	363569

12	9 or 10 or 11	364085

13	8 and 12	1425

14	debias.mp.	52

15	debiasing strategy.mp.	13

16	medical education.mp. or exp Education, Medical/	21855

17	educational initiative.mp.	121

18	bias mitigation.mp.	18

19	debiasing.mp.	346

20	14 or 15 or 16 or 17 or 18 or 19	22352

21	13 and 20	28

22	limit 21 to ("review" or "scientific integrity review" or "systematic review")	0

23	21 not 22	28

Ovid Healthstar <1966 to March 2022>

1	Cognitive bias.mp.	996

2	premature closure.mp.	426

3	exp Heuristics/ or heuristics.mp.	2407

4	exp Bias/ or anchoring bias.mp.	67213

5	overconfidence.mp.	370

6	fallacy.mp.	1146

7	exp Prejudice/ or prejudice.mp.	33589

8	1 or 2 or 3 or 4 or 5 or 6 or 7	105232

9	diagnostic error.mp. or exp Diagnostic Errors/	100430

10	misdiagnosis.mp.	9660

11	exp Delayed Diagnosis/ or exp Diagnosis/ or diagnosis.mp. or exp Missed Diagnosis/	6267940

12	9 or 10 or 11	6268699

13	8 and 12	55978

14	debias.mp.	15

15	debiasing strategy.mp.	6

16	medical education.mp. or exp Education, Medical/	179162

17	educational initiative.mp.	316

18	bias mitigation.mp.	18

19	debiasing.mp.	111

20	14 or 15 or 16 or 17 or 18 or 19	179506

21	13 and 20	936

22	limit 21 to ("review" or "scientific integrity review" or "systematic review")	49

23	21 not 22	887
